# Characterization of Long COVID Definitions and Clinical Coding Practices

**DOI:** 10.1101/2023.10.04.23296301

**Authors:** Monika Maripuri, Andrew T Dey, Jacqueline P Honerlaw, Chuan Hong, Yuk-Lam Ho, Vidisha Tanukonda, Alicia W Chen, Vidul Ayakulangara Panickan, Xuan Wang, Harrison G Zhang, Doris Yang, Malarkodi Jebathilagam Samayamuthu, Michele Morris, Shyam Visweswaran, Brendin R Beaulieu-Jones, Rachel B Ramoni, Sumitra Muralidhar, Michael J. Gaziano, Katherine P Liao, Zongqi Xia, Gabriel A Brat, Tianxi Cai, Kelly Cho

## Abstract

**Background:** Long COVID characterized as post-acute sequelae of SARS-CoV-2 (PASC) has no universal clinical case definition. Recent efforts have focused on understanding long COVID symptoms and electronic health records (EHR) data provides a unique resource for understanding this condition. The introduction of the International Classification of Diseases (ICD)-10 code U09.9 for “Post COVID-19 condition, unspecified” to identify patients with long COVID has provided a method of evaluating this condition in EHRs, however, the accuracy of this code is unclear.

**Objective:** Our study aimed to characterize the utility and accuracy of the U09.9 code across three healthcare systems - The Veterans Health Administration (VHA), Beth Israel Deaconess Medical Center (BIDMC) and The University of Pittsburgh Medical Center (UPMC) against patients identified with long COVID via a chart review by operationalizing the World Health Organization (WHO) and Centers for Disease Control (CDC) definitions.

**Methods:** COVID positive patients with either a U07.1 ICD code or positive polymerase chain reaction (PCR) test within these healthcare systems were identified for chart review. Among this cohort we sampled patients based on two approaches i) with a U09.9 code and ii) without a U09.9 code but with a new onset PASC related ICD code, which allows us to assess the sensitivity of the U09.9 code. To operationalize the long COVID definition based on health agency guidelines, we grouped symptoms into a “core” cluster of 11 commonly reported symptoms among long COVID patients and an extended cluster, that captured all other symptoms by disease domain.

Patients having at least 2 symptoms persisting for >=60 days that were new onset after their COVID infection, with at least one symptom in the core cluster, were labeled as having long COVID per chart review. We compared the performance of the code across three health systems and across different time periods of the pandemic.

**Results:** A total of 900 patient charts were reviewed across 3 healthcare systems. The prevalence of long COVID among the cohort with the U09.9 ICD code, based on the operationalized WHO definition was between 23.2%-62.4% across these healthcare systems. We also evaluated a less stringent version of the WHO definition and the Centers for Disease Control (CDC) definition and observed an increase in the prevalence of long COVID at all three healthcare systems.

**Conclusions:** This is one of the first studies to evaluate the U09.9 code against a clinical case definition for long COVID, as well as the first to apply this definition to EHR data using a chart review approach on a nationwide cohort across multiple healthcare systems. This chart review approach can be implemented at other EHR systems to further evaluate the utility and performance of the U09.9 code.

## Introduction

Characterizing the public health burden of post-acute sequalae of SARS-CoV-2 (PASC), also known as long COVID, has been difficult given that multiple clinical case definitions have been proposed by various international health agencies. [1-3] While the exact components of these definitions vary, they share some common underlying features such as the development of symptoms that are new onset after a COVID-19 infection and persistence of new onset symptoms for a duration of time post-acute infection period. Electronic health records (EHR) provide a uniquely rich resource for studying this condition at scale and there have been multiple efforts to describe long-COVID symptoms and estimate prevalence in various EHR systems. [4-9]

Specifically, the introduction of the International Classification of Diseases (ICD)-10 code U09.9 for “Post COVID-19 condition, unspecified” has provided an alternative method of evaluating this condition in EHRs, and its use has been described in various healthcare systems. [10-13] However, the accuracy of the U09.9 code in identifying long COVID has not yet been evaluated against any existing clinical case definitions in a multicenter setting. Clinical coding of long COVID has the potential for misclassification given the heterogeneity and ambiguity around the definition of Long COVID. [14]

Our study aims were to i) Characterize the use of ICD-10 code U09.9 across three healthcare systems and ii) evaluate the accuracy of the U09.9 code against patients identified with long COVID via chart review.

## Methods

### Data Sources and Study Cohort

The Consortium for Clinical Characterization of COVID-19 by EHR (4CE) is an international consortium for data driven studies on the COVID-19 pandemic. [15] Three health systems from the 4CE Consortium contributed chart review results for the current study, namely the national Veterans Health Administration (VHA), Beth Israel Deaconess Medical Center (BIDMC) and University of Pittsburgh Medical Center (UPMC). Over 15 million patients are collectively provided care across all three health systems. [16-18] The VHA is the largest integrated healthcare system in the United States, with 171 medical centers throughout the country. [16] BIDMC is an academic medical center that is a part of Beth Israel Lahey healthcare system located in Boston and UPMC is a Pittsburgh based healthcare system with 40 hospitals across Pennsylvania. [17-18] All activities were approved by the Institutional Review Boards (IRB) at each of the participating healthcare systems.

We used EHR data from the three health systems to identify COVID positive patients, define patient characteristics and obtain clinical notes for chart review. The sampling strategy for our chart review is described in Figure 1. Patients who had their first incidence of COVID-19 diagnosis reported within the participating health systems EHR with either a U07.1 ICD code for “COVID-19”, or a positive polymerase chain reaction (PCR) test performed between March 1^st^, 2020, to December 31^st^, 2021, were identified for chart review. From this COVID positive cohort, we then sampled patients based on two approaches: i) the presence of the U09.9 ICD code, which was first introduced in the United States in October 2021 or ii) the presence of at least one new onset PASC related ICD code if the patient did not have a U09.9 code. These PASC related ICD codes were selected to enrich the chart review sample for patients who may potentially have long COVID. At the VHA we further sampled patients from two time periods, those who were COVID positive before September 1, 2021 (pre-U09.9 period) and those who were COVID positive after this date (post-U09.9 period).

**Figure 1.**
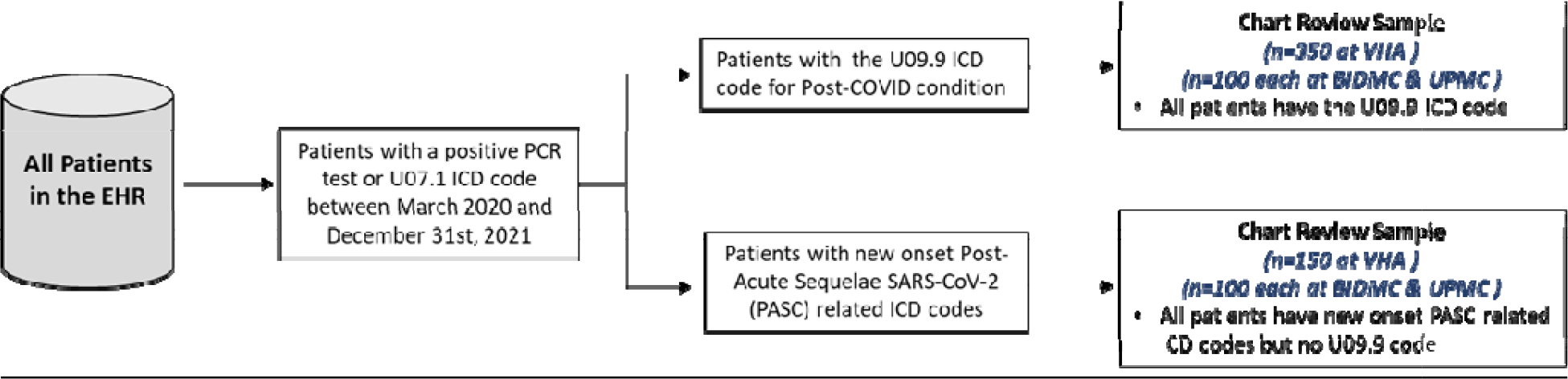
Patient sampling strategy for chart review. EHR-Electronic Health Records, ICD-International Classification of Diseases, PCR - Polymerase Chain Reaction, PASC – Post Acute Sequelae SARS-CoV-2, VHA – Veterans Health Administration, BIDMC – Beth Israel Deaconess Medical Center, UPMC – University of Pittsburgh Medical Center

The presence of PASC related feature ICD codes were identified via a data-driven process using EHR data from ten healthcare systems at 4CE. [19] Initial steps consisted of extracting longitudinal codified features such as ICD codes and mapping these codified features to Phecodes for new onset of conditions after COVID-19 infection. [20] New onset conditions were defined as those which were not present before initial COVID-19 infection. The conditions were selected such that patients with COVID-19 are associated with higher risk of a new onset of the condition after adjusting for baseline confounders such as age, sex, self-reported race, and healthcare utilization. (Supplement 1)

### Chart Review Approach

The primary aim of our chart review was to operationalize the clinical case definition for Long COVID by the World Health Organization (WHO), with secondary aims to compare against a less stringent WHO definition and the Centers for Disease Control (CDC) definition.[3,21] The chart review protocol (Supplement 2) was developed at the VHA with guidance from 4CE Consortium subject matter experts to operationalize the WHO and CDC clinical case definitions. Long COVID symptoms were identified from the WHO definition as well as through a literature review, and 11 commonly occurring symptoms among long COVID patients were classified into a “core” symptom cluster. [22-25] All other symptoms were classified into an “extended” symptom cluster based on their disease domain, which included cardiovascular, neurological, dermatological, musculoskeletal, digestive, and respiratory domains. For patients to be labeled as having long COVID per the WHO definition during the chart review (reported here as “WHO-2”), at least 2 new onset symptoms after their COVID infection were required (Figure 2). These could be either i) two “core” symptoms or ii) one “core” and one “extended” cluster symptom, each of which must have persisted for 60 days or longer. All sampled patient charts had at least 6 months of clinical notes for review after the incident COVID-19 infection to allow appropriate assessment of symptoms. During chart review, all symptoms were collected based on their onset and duration of persistence for either 30 or 60 days (Supplement 2) to allow evaluation against multiple long COVID definitions. The less stringent WHO definition (reported here as “WHO-1”) was defined as a patient having just one core symptom persisting for at least 60 days or longer and the CDC definition was defined as a patient having just one core symptom persisting for at least 30 days.

**Figure 2.**
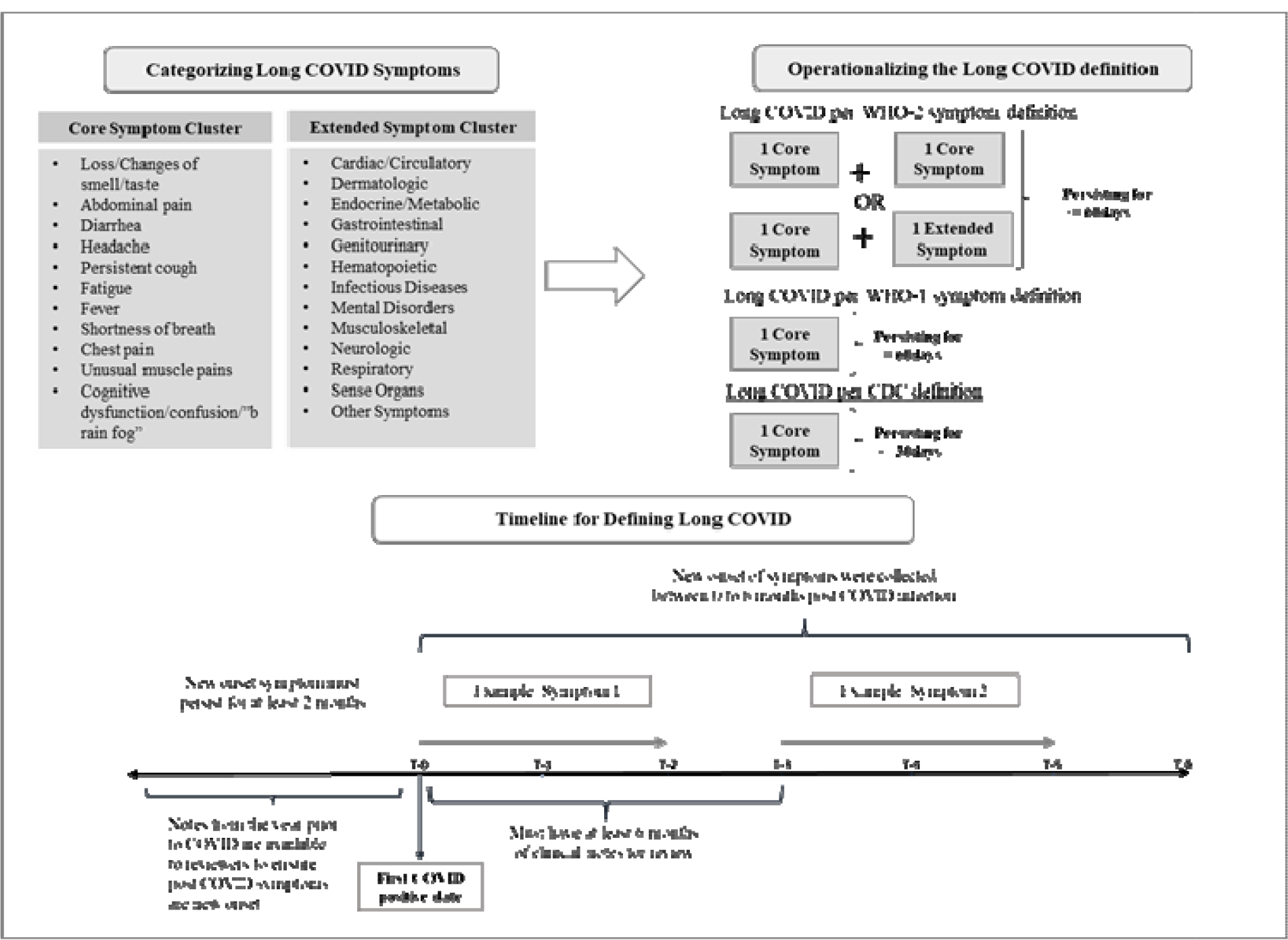
Chart Review Approach. WHO-World Health Organization, CDC-Centers for Disease Control

Reviewers had access to all clinical notes 1 year prior to the incident COVID-19 infection to determine baseline symptoms and conditions. Any symptoms present at the time of the incident COVID-19 infection or exacerbations of existing conditions were not considered new onset and thus not captured in the review. However, symptoms that waxed and waned over time were captured. At the VHA a total of 500 patient charts were reviewed and 200 patient charts were reviewed at each of the other two sites - BIDMC and UPMC.

### Characterizing U09.9 ICD Code

To characterize the use of the U09.9 ICD code in clinical practice, we investigated the following three metrics i) the frequency of the U09.9 code used over time from October 2021 to September 2022; ii) the frequency of the U09.9 code use across Veterans Integrated Service Networks (VISNs, which are regional systems of care at the VHA); and iii) the time elapsed between COVID diagnosis and U09.9 code assignment.

## Results

### Characteristics of Study Cohort

Demographics across the health systems varied notably, at the VHA patients who had a COVID-19 diagnosis between March 1, 2020, and December 31, 2021, were generally White and male veterans. Among those who were assigned a U09.9 code the demographics were generally similar with a few notable exceptions (Table 1). At BIDMC and UPMC however the demographics were different with a higher proportion of females who were assigned the U09.9 code. We also observed across all three health systems that a higher proportion of those assigned a U09.9 code had received at least one dose of a COVID vaccine.

**Table 1.** Patient Demographics. VHA – Veterans Health Administration, BIDMC - Beth Israel Deaconess Medical Center, UPMC - University of Pittsburgh Medical Center.

|  | *All COVID positive\* patients from March 1st, 2020, to December 31st, 2021* |  |  | *All COVID positive\* patients with a U09.9 ICD code* |  |  | *All COVID positive\* patients with a new onset PASC feature* |  |  |
| --- | --- | --- | --- | --- | --- | --- | --- | --- | --- |
| *Health Systems* | *VHA* | *BIDMC* | *UPMC* | *VHA* | *BIDMC* | *UPMC* | *VHA* | *BIDMC* | *UPMC* |
| **Total** | **307,909** | **30,294** | **147,653** | **22,196** | **164** | **6,057** | **294,302** | **7,245** | **92,120** |
| Age at incident COVID Diagnosis - Mean (SD) | 59.2 (16.1) | 47.5 (20.5) | 45.7 (23.8) | 61.7 (15.1) | 54.7 (13.9) | 55.1 (17.2) | 61.2 (15.7) | 54.6 (18.4) | 47.5 (24.1) |
| **Sex** |  |  |  |  |  |  |  |  |  |
| Male | 272,957 (88.7%) | 16,579 (54.7%) | 63,421 (43%) | 19,275 (86.8%) | 58 (35.4%) | 2219 (36.6%) | 260,055 (88.4%) | 2,947 (40.67%) | 38,348 (41.6%) |
| Female | 34,898 (11.3%) | 13,715 (45.3%) | 84,232 (57%) | 2,921 (13.2%) | 106 (64.6%) | 3838 (63.4%) | 34,214 (11.6%) | 4,298 (59.3%) | 53,772 (58.4%) |
| **Race** |  |  |  |  |  |  |  |  |  |
| White | 208,457 (67.7%) | 8,983 (29.7%) | 125,503 (85%) | 16,102 (72.5%) | 105 (64%) | 4,965 (82%) | 197,237 (67.0%) | 3,096 (42.7%) | 77,471 (84.1%) |
| Black or African American | 69,067 (22.4%) | 5,074 (16.7%) | 14,966 (10.1%) | 3,621 (16.3%) | 29 (17.7%) | 880 (14.5%) | 69,037 (26.5%) | 1,730 (23.9%) | 10,564 (11.5%) |
| Asian | 3,326 (1.1%) | 1,200 (4%) | 1,303 (0.9%) | 249 (1.1%) | 4 (2.4%) | 70 (1.2%) | 3,360 (1.1%) | 270 (3.7%) | 986 (1.1%) |
| American Indian or Alaska Native | 3,521 (1.1%) | 31 (0.1%) | 704 (0.5%) | 273 (1.2%) | 1 (0.6%) | 0 (0%) | 3,282 (1.1%) | 8 (0.1%) | 493 (0.5%) |
| Native Hawaiian or Pacific Islander | 3,189 (1%) | 21 (0.07%) | 35 (0.02%) | 239 (1.1%) | 0 (0%) | 0 (0%) | 3,018 (1.0%) | 9 (0.1%) | 0 (0%) |
| Not Reported | 0 (0%) | 14,985 (49.5%) | 5,141 (3.5%) | 0 (0%) | 25 (15.2%) | 142 (2.3%) | 0 (0%) | 2,132 (29.4%) | 2,606 (2.8%) |
| **Vaccination Status** |  |  |  |  |  |  |  |  |  |
| Received at least one dose of COVID vaccine | 199,235 (64.7%) | 9,649 (31.9%) | 58,244 (39.4%) | 15,295 (68.9%) | 112 (68.3%) | 3,521 (58.1%) | 203,569 (69.2%) | 4,470 (61.7%) | 41,271 (44.8%) |
| Did not receive at least one dose of COVID vaccine | 104,907 (34.1%) | 20,645 (68.1%) | 89,409 (60.6%) | 6,684 (30.9%) | 52 (31.7%) | 2,536 (41.9%) | 87,569 (29.8%) | 2,775 (38.3%) | 50,849 (55.2%) |

*\*COVID positive is defined as a patient having either a U07.1 ICD-10 Code or a documented Positive PCR Test*

We observed a substantial variation in the use of U09.9 code to diagnose long COVID over time and region. Figures 3 and 4 show the results of our characterizations of the U09.9 code use 12 months following its introduction in the Unites States, on October 1, 2021. Figure 3 shows the frequency of the U09.9 code diagnosis per 10,000 new COVID cases that occurred in the last 12 months from when they received the code. Frequency of the U09.9 code used to diagnose long COVID was highest from January to March 2022 at health system 1, February to Mach 2022 at health system 2 and December 2021 to January 2022 at health system 3.

**Figure 3.**
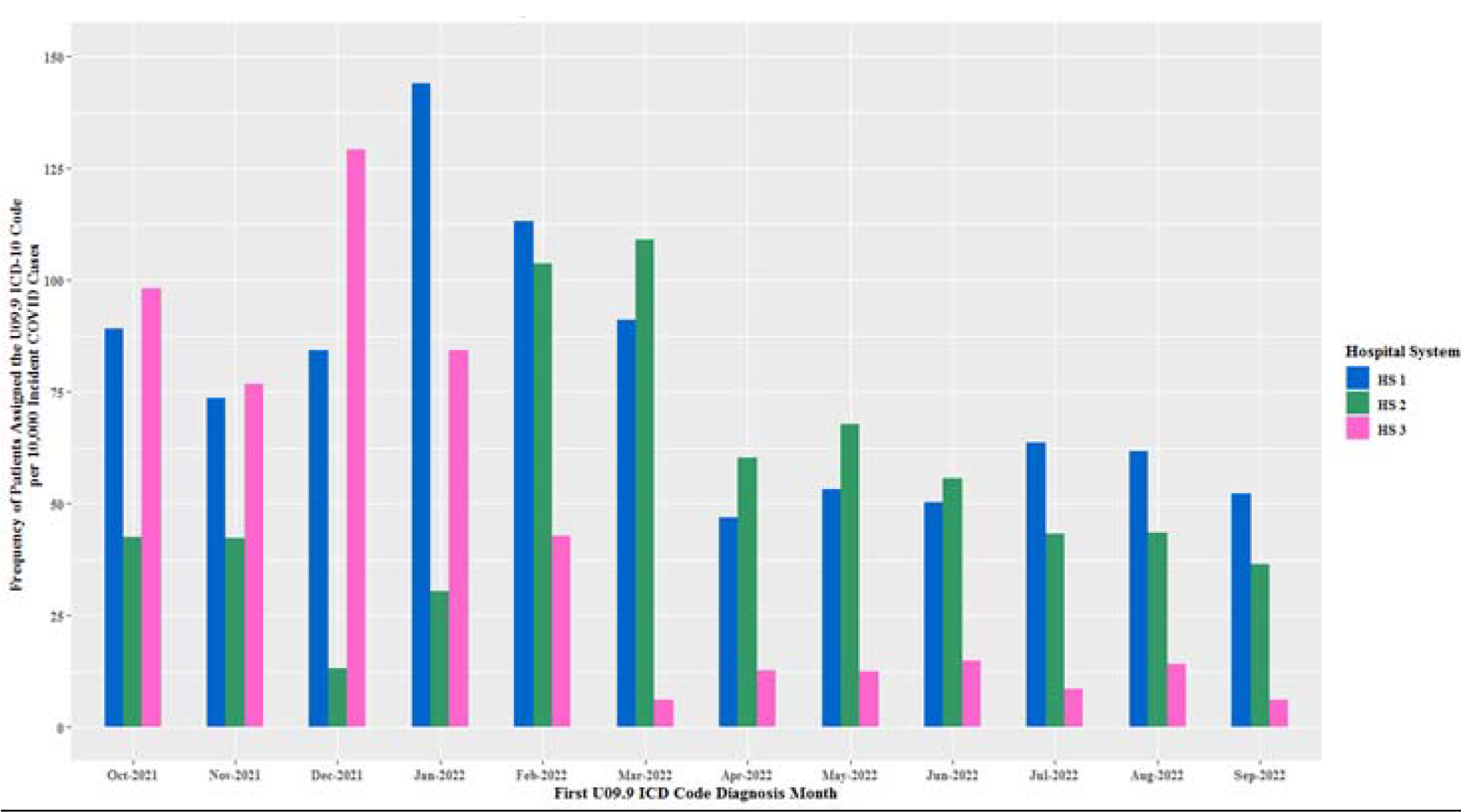
Frequency of a new U09.9 ICD code assignment per 10,000 new COVID cases that occurred within the previous 12 months.

**Figure 4.**
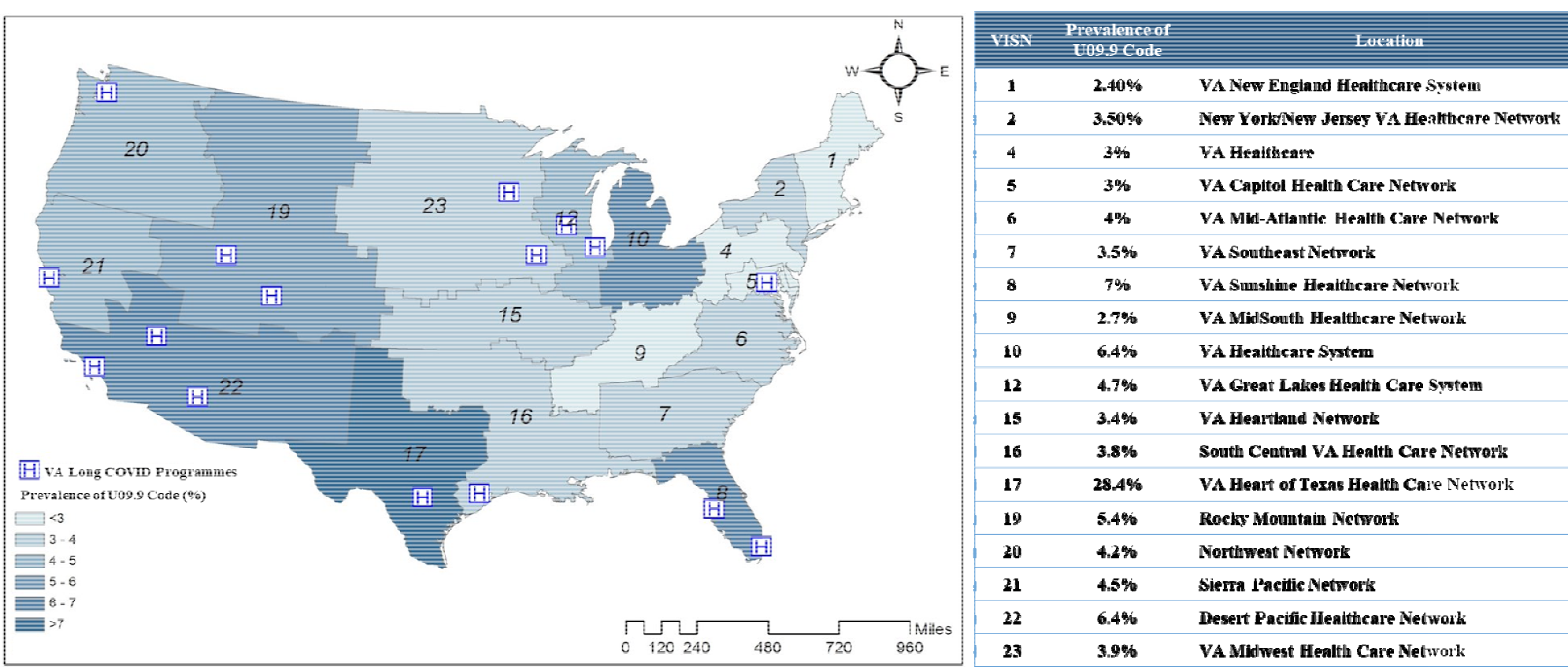
Prevalence of the U09.9 ICD code by region - VA Veterans Integrated Service Networks (VISN)

There were also large regional differences in the use of U09.9 code across VHA Healthcare system VISNs (Figure 4). VISN 17 assigned the U09.9 code to 28.4% of all patients who received this code at the VHA, while VISN1 assigned the U09.9 code to just 2.4% of all patients who received the code at the VHA.

### Chart Review Results

Chart review at the VHA was conducted by 2 clinical reviewers (M.M and J.H.) with a 1% overlap and an inter-rater reliability of 80%. At BIDMC and UPMC chart review was conducted by 1 clinical reviewer (B.B and M.S) respectively. The most common symptoms identified during chart review among patients with long COVID as per the WHO-2 definition were shortness of breath, fatigue, cough, and loss of smell or taste from the core symptom cluster. Among the extended symptom clusters, we most commonly saw symptoms across the cardiovascular, gastrointestinal, neurological, and respiratory disease domains.

Chart review was performed on a total of 900 patients infected with COVID-19 across three healthcare systems. We anonymized these three institutions to provide an unbiased interpretation of the results. The positive predictive value (PPV) of long COVID from chart review, among the sample of patients with the U09.9 ICD code per the WHO-2 definition was 29.8% at health system 1 and 62.4%, 23.2% at health system 2 and 3 respectively. (Figure 5) Among the sample of patients with a new onset PASC features the PPV of long COVID was 7% at health system 1 and 6.74%, 3% at health system 2 and 3 respectively. However, when we consider the WHO-1 and the CDC definitions, the PPV of long COVID was higher at all three health systems but at health system 2 the PPV was slightly higher for the WHO-1 definition but remained the same for the CDC definition. (Figure 5) The overall performance of the U09.9 code based on the WHO-2 definition resulted in a weighted sensitivity of 15% at health system 1 and 4.9%, 19.1% at health system 2,3 respectively.

**Figure 5.**
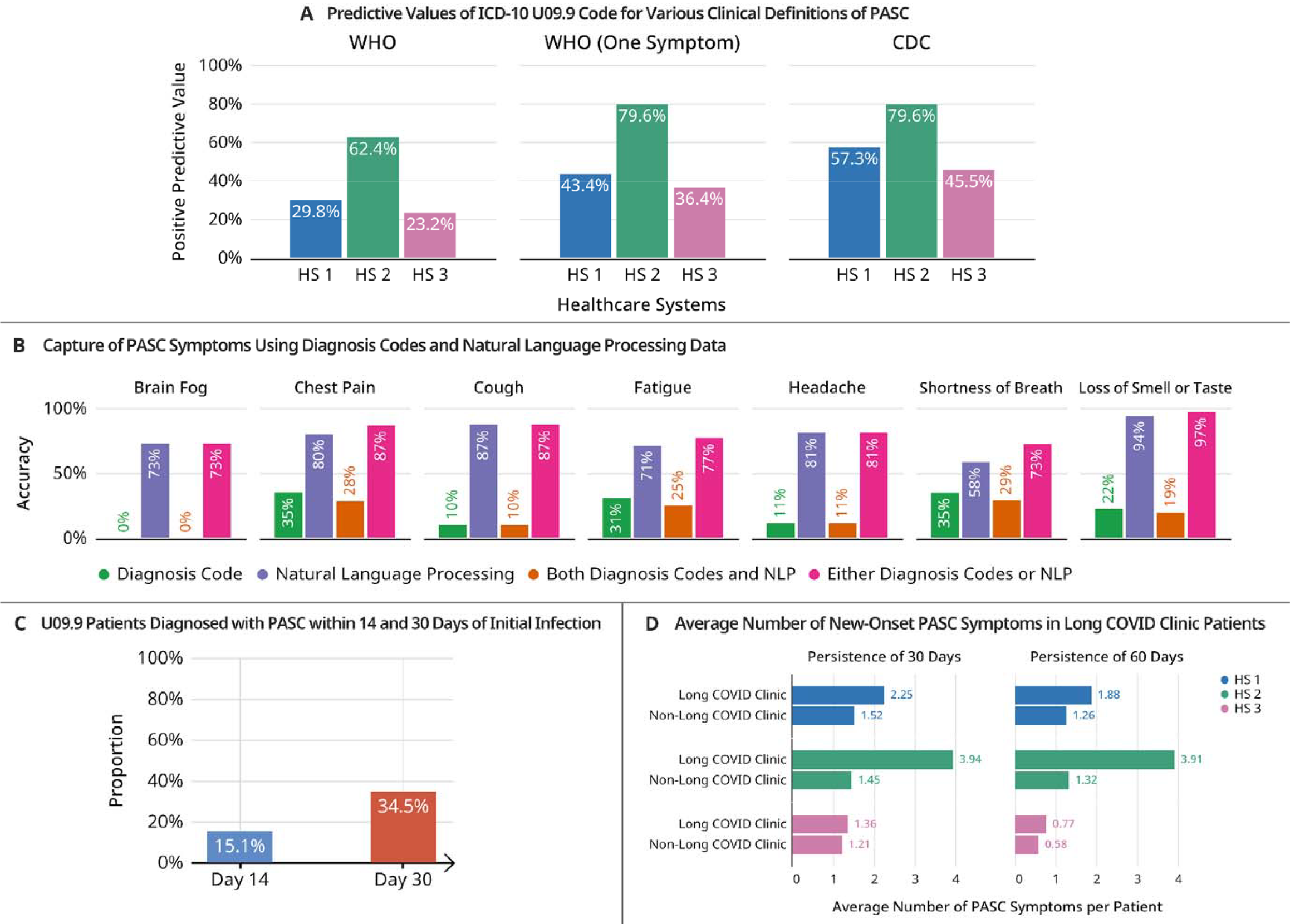
Comparison of results among all sites. ICD – International Classification of Diseases, WHO-World Health Organization, CDC-Centers for Disease Control, PASC-Post Acute Sequelae of SARS-CoV-2

Additionally, at the VHA we looked at the prevalence of long COVID among patients with the U09.9 ICD code across the different time periods based on their first COVID infection date. From the chart reviewed cohort 44.64% had long COVID from the pre-U09.9 period and 22.75% from the post-U09.9 period at the VHA. The positive predictive value (PPV) of patients with long COVID in the pre-U09.9 period is higher as many of these patients were back coded.

Through our chart review we observed that patients were given the U09.9 code over a wide range of time from less than 29 days to over 365 days following their initial COVID-19 infection. Most patients did not have persisting symptoms post-acute infection and waxing - waning of symptoms was frequently observed.

## Discussion

### Principal Results

This study provides a comprehensive, multicenter evaluation of the U09.9 code against proposed clinical case definitions for long COVID. It is also the one of the first studies to apply a clinical case definition to EHR data using a chart review approach. The use of EHR data allowed evaluation of the U09.9 code across multiple healthcare systems nationwide. The availability of ample clinical notes enabled reviewers to ascertain whether observed symptoms post COVID infection were truly new onset and to evaluate the duration of new symptoms, which are critical components of case definitions for long COVID. Another strength of our study was that we evaluated both the WHO and CDC clinical case definitions for long COVID since one universal definition is not currently available. Our symptom collection approach (Supplement 2) captured discrete symptoms by duration of 30 or 60 days, which allowed for multiple case definitions to be applied.

There were large variations in the accuracy of use of the U09.9 code for long COVID. We observed that one center had a much higher predictive value for patients with long COVID among the U09.9 cohort across all three definitions than the other two healthcare systems. This health system also had the highest average number of new onset symptoms among patients seen in long COVID clinics. The U09.9 code assignment at this health system could have been more accurate due to a higher proportion of patients being seen at long COVID clinics.

While looking at the capture of PASC symptoms using ICD codes and Natural Language Processing (NLP) data, the performance of NLP is significantly better for all the commonly occurring long COVID symptoms (Figure 5). [14] The accuracy of using either a diagnosis code or NLP had the best results with 97% accuracy for loss of smell or taste and 87% accuracy for chest pain and cough.

### Limitations

There were several limitations with our study. The cohort at the VHA had a higher proportion of male patients who were generally older, and predominantly white. Incident COVID-19 infection was required for inclusion in the chart review, and it is possible that patients had an infection outside of the health systems which was not recorded in the EHR. Patients may have also had symptoms that were not reported at healthcare system visits. The regional variation in the number of long COVID clinics may have led to differential capture of symptoms for patients seen at long COVID clinics versus those seen by other care providers. We observed that symptoms among these patients were well documented as most long COVID clinics have a specific template for evaluating and capturing of COVID-19 symptoms. [26] In some instances, it was difficult to assess whether a symptom was truly new onset due to COVID-19 infection or a result of underlying health conditions noted at baseline. While the WHO definition has been in use since 2021, long COVID is still an evolving disease, and the case definition may change over time as the condition is further characterized. We also faced some challenges in optimizing a heterogenous and sparse data capture within the EHR systems.

### Comparison with Prior Work

The use of the U09.9 code has been described in several cohorts. The NIH’s National COVID Cohort Collaborative (N3C) reported on the growing use of U09.9 from October 2021 through January 2022 in a nationwide cohort of 21,072 patients with the code [10]. However, N3C did not require patients in the cohort to have a positive COVID test to evaluate the use of the U09.9 code and 32.3% of patients did not have a COVID index date. McGrath and colleagues also reported increasing use of the U09.9 code in the months following its release in the nationwide Health Verity cohort of 56,143 patients with a U09.9 code which included children under 18 [12].

Similar to N3C, this cohort did not require a COVID positive test for evaluation of the U09.9 code and only 70.4% had a documented COVID infection. Of the COVID positive patients with U09.9, the median time from infection to U09.9 diagnosis was 56 days. A study in Sweden by Bygdell et al reported 10,196 patients with the U09.9 code, [11] They also found that 2.0% of the COVID positive population in the two largest regions of Sweden had U09.9 at least 28 days post infection.

## Conclusions

Our findings suggest that the U09.9 code should be used judiciously in EHR-based studies of long COVID. Given the low PPV of the U09.9 code, it’s usage as a proxy for long COVID is not recommended. However, the sensitivity of the code makes it useful for identifying patients who may have long COVID and thus require further clinical evaluation.

This was one of the initial efforts towards validating long COVID against a clinical case definition and the U09.9 code through a chart review on a nationwide cohort. The chart review approach developed at the VA can be implemented at other EHR systems to further evaluate the utility and performance of the U09.9 code. Further efforts to develop a more refined and reproducible phenotyping algorithm for long COVID is underway utilizing the chart review labels from our study for algorithm training and development.

## Data Availability

Data cannot be shared publicly because of policies regarding data privacy and security. All relevant summary level data are included in the manuscript.

