## Supplement 1 for "Characterization of Long COVID Definitions and Clinical Coding Practices"

### Supplement 1 – Selected PASC feature Phecodes

Marginal testing using a logistic regression framework was then performed to identify associated new onset conditions emerging three months after initial infection. Conditions which passed marginal testing were then subject to conditional randomization analyses via distillation to robustly test whether a condition’s new onset is conditionally dependent on prior COVID-19 infection.[19] The Benjamini Hochberg procedure was used to adjust for multiple comparisons. [20]

| **Phecode** | **Description** |
| --- | --- |
| 38 | Septicemia |
| 38.1 | Gram negative septicemia |
| 41.9 | Infection with drug-resistant microorganisms |
| 54 | Herpes simplex |
| 79 | Viral infection |
| 79.9 | Viremia, NOS |
| 136 | Other infectious and parasitic diseases |
| 250.2 | Type 2 diabetes |
| 250.23 | Type 2 diabetes with ophthalmic manifestations |
| 250.41 | Impaired fasting glucose |
| 250.42 | Other abnormal glucose |
| 250.7 | Diabetic retinopathy |
| 257.1 | Testicular hypofunction |
| 261.2 | Vitamin B-complex deficiencies |
| 261.4 | Vitamin D deficiency |
| 271.3 | Intestinal disaccharidase deficiencies and disaccharide malabsorption |
| 274.1 | Gout |
| 276.11 | Hyperosmolality and/or hypernatremia |
| 278.1 | Obesity |
| 278.11 | Morbid obesity |
| 278.4 | Abnormal weight gain |
| 285.21 | Anemia in chronic kidney disease |
| 290.1 | Dementias |
| 290.11 | Alzheimer's disease |
| 290.16 | Vascular dementia |
| 292 | Neurological disorders |
| 292.1 | Aphasia/speech disturbance |
| 292.3 | Memory loss |
| 292.4 | Altered mental status |
| 327 | Sleep disorders |
| 327.3 | Sleep apnea |
| 339 | Other headache syndromes |
| 345 | Epilepsy, recurrent seizures, convulsions |
| 348.8 | Encephalopathy, not elsewhere classified |
| 349 | Other and unspecified disorders of the nervous system |
| 380.4 | Impacted cerumen |
| 386.9 | Dizziness and giddiness (Light-headedness and vertigo) |
| 389.4 | Tinnitus |
| 395.3 | Nonrheumatic tricuspid valve disorders |
| 401.1 | Essential hypertension |
| 401.22 | Hypertensive chronic kidney disease |
| 415 | Pulmonary heart disease |
| 415.11 | Pulmonary embolism and infarction, acute |
| 418 | Nonspecific chest pain |
| 427.9 | Palpitations |
| 433.8 | Late effects of cerebrovascular disease |
| 452 | Other venous embolism and thrombosis |
| 452.2 | Deep vein thrombosis [DVT] |
| 464 | Acute sinusitis |
| 465 | Acute upper respiratory infections of multiple or unspecified sites |
| 476 | Allergic rhinitis |
| 477 | Epistaxis or throat hemorrhage |
| 480 | Pneumonia |
| 480.2 | Viral pneumonia |
| 483 | Acute bronchitis and bronchiolitis |
| 501 | Pneumonitis due to inhalation of food or vomitus |
| 509.1 | Respiratory failure |
| 509.8 | Dependence on respirator [Ventilator] or supplemental oxygen |
| 510 | Other diseases of lung |
| 512.7 | Shortness of breath |
| 512.8 | Cough |
| 512.9 | Other dyspnea |
| 519.8 | Other diseases of respiratory system, NEC |
| 573.7 | Abnormal results of function study of liver |
| 573.9 | Abnormal serum enzyme levels |
| 585.31 | Renal dialysis |
| 585.32 | End stage renal disease |
| 591 | Urinary tract infection |
| 592.11 | Acute cystitis |
| 605 | Erectile dysfunction [ED] |
| 614.52 | Vaginitis and vulvovaginitis |
| 619 | Noninflammatory female genital disorders |
| 619.4 | Noninflammatory disorders of vagina |
| 626 | Disorders of menstruation and other abnormal bleeding from female genital tract |
| 704.1 | Alopecia |
| 707.1 | Decubitus ulcer |
| 726.3 | Bursitis |
| 728.7 | Fasciitis |
| 729 | Other disorders of soft tissues |
| 731 | Osteitis deformans and osteopathies associated with other disorders classified elsewhere |
| 755.1 | Congenital deformities of feet |
| 771.1 | Swelling of limb |
| 772.3 | Muscle weakness |
| 783 | Fever of unknown origin |
| 790 | Nonspecific findings on examination of blood |
| 790.6 | Other abnormal blood chemistry |
| 792.1 | Papanicolaou smear of cervix or vagina with atypical squamous cells |
| 798 | Malaise and fatigue |
| 798.1 | Chronic fatigue syndrome |
| 994.2 | Sepsis |

**ICD to Phecodes mapping can be found on the* [*PheWAS Catalog*](https://phewascatalog.org/)
