## Supplement 2 for "Characterization of Long COVID Definitions and Clinical Coding Practices"

### Supplement 2 – Chart Review Protocol

##### Goals

1. **Primary Goal**
   a. SPECIFICITY OF CODES. How well do coding filters identify patients with a clinically meaningful definition of COVID-19.
   b. NEW ONSET. Did the patient have any of these health conditions prior to the diagnosis of COVID-19?
2. **Secondary Goal**
   a. What constellation of associated symptoms are consistent with U09.9 and (stop codes for clinic)?

##### Definition of Long COVID-19

The definition of long covid for this chart review is drawn from the [WHO consensus definition 2021](https://www.who.int/publications/i/item/WHO-2019-nCoV-Post_COVID-19_condition-Clinical_case_definition-2021.1):

*“Post COVID-19 condition occurs in individuals with* ***a history of probable or confirmed SARS-CoV-2*** *infection,* ***usually 3 months from the onset of COVID-19 with symptoms that last for at least 2 months and cannot be explained by an alternative diagnosis.*** *Common symptoms include* ***fatigue, shortness of breath, cognitive dysfunction*** *but also others (see* ***Table 3*** *and* ***Annex 2****) which generally have an* ***impact on everyday functioning.*** *Symptoms may be* ***new onset,*** *following initial recovery from an acute COVID- 19 episode, or* ***persist*** *from the initial illness. Symptoms may also* ***fluctuate*** *or* ***relapse*** *over time. A separate definition may be applicable for children.”*

The following operational definition was adapted from WHO to classify a patient with long COVID-19 for this chart review:

1. Symptom/pathology must be new onset, occurring after COVID-19 diagnosis.
2. Symptom/pathology must not be an exacerbation of an existing objective pre-COVID pathology.
3. Must persist for at least 2 months.
4. Must start within the 6 months after initial infection.
5. Review chart from patient who is at least 3 months post COVID-19 infection.
6. Must include at least 1 symptom/pathology from the CORE set.
7. IF only 1 CORE symptom/pathology present, must include at least 1 symptom/disease from EXTENDED set.

##### Data Sources for Review

| **ICD-10 Codes** | All available ICD codes and dates (including U09.9) |
| --- | --- |
| **Labs** | List of all positive COVID-19 tests and date |
| **Patient Notes** | All notes available in data pull timeframe |
| **COVID-19 related hospitalizations** | Admission code, date, discharge date |

**Search Protocol**

Based on initial review of first 40 charts:

- Must include at least 1 symptom/pathology from the CORE set that persisted for longer than 30/60 days.
- The symptom may be differentially captured during the 60-day period in patient notes.
  - For example, the notes may state that the patient denies SOB, then confirms SOB, then denies.
  - If the majority of notes mark the symptom as present, then the symptom should be captured in the chart review.
- Capture the timeline for core symptoms - if it’s 30 days or 60 days, to enable flexibility of data analysis

##### Reviewer Classifications

##### COVID Diagnosis and Baseline Conditions

| **Field** | **Description** | **Data entered** |
| --- | --- | --- |
| First Positive COVID Test | Date of first positive covid-19 lab test. This may be found in the structured lab data or in free text (if performed outside of health system) | mm/dd/yyyy* |

##### COVID Vaccination

| **Field** | **Description** | **Data** |
| --- | --- | --- |
| Vaccination status | Was the patient ever vaccinated? | - Yes - No |
| Vaccination date | Date of first vaccination: | mm/dd/yyyy* |

##### COVID-19 Related Symptom(s) and Duration

| **Field** | **Description** | **Data** |
| --- | --- | --- |
| CORE Symptom Cluster | Specific, very common symptoms that are nearly always present.  Only capture:   - Symptoms with onset after COVID-19 infection - Symptoms that start within 6 months of covid infection - Symptoms that persist for at least 30 or 60 days | **Check all that apply** \| **Start Date*** *(mm/dd/yyyy)*; Loss/Changes of smell/taste \|; Abdominal pain \|; Diarrhea \|; Headache \|; Persistent cough \|; Fatigue \|; Fever \|; Shortness of breath \|; Chest pain \|; Unusual muscle pains \|; Cognitive dysfunction/confusion/”fog” \| |
| EXTENDED Symptom Cluster | Manual grouping of symptoms for ease of chart review. Based on phecode groups.  Only capture:   - Symptoms with onset after COVID-19 infection - Symptoms that start within 6 months of covid infection - Symptoms that persist for at least 30 or 60 days   Start date is for the first noted symptom in the cluster | **Check all that apply** \| **Symptom **** *(free text)* \| **Start Date*** *(mm/dd/yyyy)*; Cardiac/Circulatory \| \|; Dermatologic \| \|; Endocrine/metabolic \| \|; Gastrointestinal \| \|; Genitourinary \| \|; Hematopoietic \| \|; Infectious disease \| \|; Mental disorders \| \|; Musculoskeletal \| \|; Neurologic \| \|; Respiratory \| \|; Sense organs \| \|; Symptoms \| \| |
| Long COVID | Does that patient meet the definition of long covid?   - At least 1 symptom marked in CORE Symptom Cluster - IF only 1 CORE symptom/pathology present, must include at least 1 symptom/disease from EXTENDED set. | - Yes - No - N/A |

**Mark “99” for unknown month or day fields*

***List each of the symptoms identified in the cluster*

##### Reviewer Comments

| **Field** | **Description** | **Data entered** |
| --- | --- | --- |
| Reviewer comment | Free text for comments | Free text |
